# The incidence and predictors of surgical site infection in a tertiary hospital in southern Ethiopia; A prospective cohort study

**DOI:** 10.1101/2025.07.19.25331828

**Authors:** Jenenu Getu Bekele, Yoseph Tafesse, Niguse Mekonnen Kara, Ashenafi Desalegn, Siyum Solomon, Tadiwos Utalo Urkashe

## Abstract

**Background:** Surgical site infection (SSI) represents a significant burden of hospital-acquired infections, particularly in developing countries, leading to prolonged hospitalizations, readmissions, long-term disability, antibiotic resistance, and increased mortality. Despite this substantial public health concern, the incidence and predictors of SSI remain understudied in southern Ethiopia.

**Methods:** An institution-based prospective cohort study was conducted at Wolaita Sodo University Comprehensive Specialized Hospital (WSUCSH) from August 1 to November 30, 2024. A total of 261 adult surgical patients were followed from admission until 30 days postop. The data were analyzed via STATA Version 14. The cumulative incidence of SSI and time to event were estimated via Kaplan Meier (KM) survival analysis, and the log-rank test was used to compare differences in time to SSI among groups. Cox proportional hazards regression was then employed to identify predictors of SSI. Adjusted hazard ratios (AHR) with 95% confidence intervals were reported, and statistical significance was set at p < 0.05.

**Results:** Among the 261 surgical patients, the cumulative incidence of SSI was 37.5% (98 cases), with an incidence density of 17.5 per 1000 person-days. Superficial SSI was the most common type (69.4%). Multivariate Cox regression analysis identified several independent predictors of SSI: underweight BMI (<18.5 kg/m^2^) [AHR=6.43; 95% CI: 3.00, 13.78], increased wound contamination (clean-contaminated [AHR=5.14; 95% CI: 1.12, 23.59], contaminated [AHR=7.53; 95% CI: 2.11, 26.53], and dirty wounds [AHR=15.51; 95% CI: 2.31, 105.83] compared with clean wounds), a lack of preoperative antibiotic prophylaxis [AHR=2.72; 95% CI: 1.17, 6.31], postoperative antibiotic prescription [AHR=5.15; 95% CI: 2.51, 10.55], and a total hospital stay duration greater than two weeks [AHR=2.80; 95% CI: 1.10, 7.10].

**Conclusion:** The incidence of surgical site infection at WSUCSH is high, with several significant contributing factors. These findings highlight the critical need for targeted interventions focused on optimizing patient nutritional status; adhering to strict infection prevention practices, including proper wound management and judicious use of antimicrobial prophylaxis; and minimizing hospital stay. The observed association between postoperative antibiotic use and increased SSI risk warrants further investigation into current antibiotic stewardship practices. Large-scale, multicenter studies are recommended to validate these findings and inform broader SSI prevention strategies in Ethiopia.

## Introduction

Surgical site infection (SSI) is defined as an infection that occurs near or at the incision site and/or deeper underlying tissue spaces and organs within 30 days of a surgical procedure or up to 90 days for implanted prosthetics (1). It remains a significant global health concern, representing one of the most common and costly hospital-acquired infections (HAIs) (2). In particular, in developing countries, the burden of SSI is amplified due to resource limitations, inadequate infection prevention practices, and a relatively high prevalence of underlying comorbidities (3). The consequences of SSI extend beyond immediate patient morbidity, contributing to prolonged hospital stays, increased rates of readmission, long-term disability, a greater risk of antibiotic resistance, substantial financial strain on already stretched healthcare systems, and a diminished quality of life for affected individuals and their families. Among its most severe forms, SSI can also lead to increased mortality (4).

The epidemiology of SSI varies considerably across different geographical regions and healthcare settings. While numerous studies have investigated the incidence and risk factors for SSI in high-income countries, data from low- and middle-income countries (LMICs), especially sub-Saharan Africa, remain relatively limited (5). According to a multicenter cohort study performed in 66 countries worldwide, 12.3% of patients will develop SSIs after surgery. The unadjusted SSI incidence varied between countries with high income (9.4%), middle income (14.0%) and low income (23.2%) (6). Globally, different studies have reported that the pooled incidence of SSI is 2.5–11% (7). In Western countries, 2 to 5% of patients who undergo clean surgery and up to 20% of patients who undergo intra-abdominal surgery will develop SSIs (8). In Africa, the cumulative incidence of SSIs ranges from 2.5 to 30.9% (9). Understanding the local context of SSI is crucial for developing targeted and effective prevention strategies. Factors such as patient demographics, prevalent comorbidities, the spectrum of surgical procedures performed, and the specific infection prevention protocols in place can significantly influence the incidence and predictors of SSI in a given setting (10).

Ethiopia, like many other developing nations, faces considerable challenges in controlling HAIs, including SSIs. According to systematic reviews and meta-analyses performed in Ethiopia, the rate of SSI ranged from 12.3% to 25.22% (11,12). Different studies conducted in different parts of Ethiopia reported conflicting results: the prevalence of SSI in Tigray was 11.5% (13), that in Dessie was 14.5% (14), that in the Amhara region was 39.1% (15), that in Jimma was 21.1% (16), that in Hawassa was 24.6% (17), that in Arbaminch was 10.5% (18), and that in Wolaita Sodo was 13% (19). While some studies have explored the prevalence of HAIs in Ethiopian hospitals, specific data on the incidence and predictors of SSI, particularly in the southern region, are scarce. This lack of localized evidence hinders the development and implementation of evidence-based interventions tailored to the unique characteristics of healthcare facilities and patient populations in this area.

The literature has established a range of factors that significantly influence the risk of surgical site infection (SSI). Patient-related risk factors include advanced age, malnutrition (e.g., low albumin), metabolic diseases (e.g., diabetes), smoking, obesity (high BMI), hypoxia, and immunosuppression. Preoperative factors such as a prolonged preoperative hospital stay, a higher American Society of Anesthesiologists (ASA) score indicating greater comorbidity burden, the presence of specific comorbidities, and suboptimal nutritional status have also been implicated. The surgery-related factors included the type and duration of the surgical procedure, the type of anesthesia administered, the appropriateness and timing of antimicrobial prophylaxis, and the degree of wound contamination (wound type). Finally, postoperative factors such as inadequate wound care practices, the use of surgical drains, and the administration of postoperative antibiotics can also impact SSI development (13–19).

Previous research in Ethiopia has explored the overall rates, causes, and costs of surgical site infections (SSIs), predominantly via retrospective and cross-sectional methodologies. A key limitation of these studies is their reliance on hospital records and follow-up limited to the point of discharge, potentially missing a significant proportion of SSIs that are either undocumented or manifest postdischarge, thus underestimating the true extent of the problem. Local observations suggest a high burden of SSI in the study area, contributing to increased morbidity and financial strain. To overcome these limitations, this prospective study aims to comprehensively assess the incidence and associated factors of SSI among surgical patients at a facility with a high case load in southern Ethiopia. Notably, this research incorporates crucial variables often overlooked in prior Ethiopian studies, including anemia/hemoglobin levels, nutritional status (albumin and BMI), and detailed intraoperative antibiotic administration (timing, type, and dose).

Therefore, this prospective cohort study was designed to investigate the incidence of surgical site infection and identify independent predictors among patients undergoing surgery at Wolaita Sodo University Comprehensive Specialized Hospital in southern Ethiopia. By prospectively following a cohort of surgical patients from admission to 30 days postdischarge, this study aims to provide robust, context-specific data that can contribute to improved patient care and inform future infection prevention efforts in the region.

## Methods

### Study setting

This prospective cohort study was conducted at Wolaita Sodo University Comprehensive Specialized Hospital (WSUCSH) in Wolaita Sodo town, southern Ethiopia. Established in 1927, WSUCSH is a teaching hospital that serves as a primary care center and a training institution for undergraduate and postgraduate medical studies. With a capacity of 350 beds, including 52 dedicated to surgical patients, the hospital caters to a catchment area of more than 3 million people. The surgical department comprises two major operating rooms, one emergency operating room, one pediatric surgery operating room, and one minor operating room. An 8-bed intensive care unit is shared with other departments. The surgical workforce includes 13 general surgeons; 5 general practitioners; and specialists in neurosurgery, urology, orthopedics, ENT, maxillofacial surgery, and pediatric surgery. The workforce is supported by 40 surgical residents, 35 anesthetists, and 70 nurses.

### Study design and period

This research employed an institution-based prospective follow-up study design. Data collection occurred over a four-month period, from August 1 to November 30, 2024.

### Population

The source population for this research was all adult patients admitted to the general surgical ward at WSUCSH. The study cohort consisted of all adult patients admitted to and operated on (both on an emergency basis and an elective basis) within the general surgery ward of WSUCSH during the defined study period.

### Inclusion and Exclusion Criteria

#### Inclusion criteria

This study included all adult patients (defined as those older than 15 years) who underwent a surgical procedure and were admitted to the General Surgery Department for that operation.

Patients were excluded if they met any of the following criteria: (1) underwent surgery at an external institution and were referred to WSUCSH for postoperative admission to the surgical ward; (2) had a documented surgical site infection present prior to the commencement of the study period; or (3) were admitted and underwent surgery specifically for the treatment of an existing surgically treatable infection.

### Sample size and sampling method

#### Sample size

For the primary objective of determining the incidence of SSI, the sample size was calculated via the single population proportion formula. This calculation incorporated a 95% confidence level, a 5% margin of error, and a previously reported proportion of SSIs in surgical patients in Ethiopia of 19.1% (20). To ensure adequate power and account for potential attrition, a 10% nonresponse rate was factored in, yielding a minimum required sample size of 261 participants.

For the second objective, the sample size was also calculated to check the adequacy of the sample size used for a survival sample size calculation power approach with the Cox proportional hazard assumption. By using the Schoenfeld formula and considering the following assumptions: significance level (α) = 0.05, power 80% (β) = 0.8, proportion of variability among covariates of interest (SD) = 0.5, square of correlation of independent variables with the outcome (R^2^) = 0.5, a clinically meaningful hazard ratio of 2.6 (reported by the effect of timely antimicrobial prophylaxis in another Ethiopian study [15]), and an event probability of 39.1% (from the same prior study). The power analysis, performed via Stata version 14.2, indicated a minimum required sample size of 194. As our study included 261 participants, the sample size is sufficient to provide adequate power for the planned survival analysis.

#### Sampling Method

A systematic random sampling approach was employed to select the study participants. The sampling interval (k) was calculated as 2 on the basis of an estimated 540 adult surgical cases expected during the 4-month study period (derived from the previous 12-month records) and a calculated sample size of 261. Owing to the continuous influx of new surgical patients throughout the data collection phase, a preexisting sampling frame was not feasible. Therefore, eligible adult general surgery patients were consecutively approached and enrolled at intervals of every two admissions until the required sample size of 261 was attained.

### Data collection tool and procedure

Data were collected via a tool adapted from the WHO ’Protocol for Surgical Site Infection Surveillance with a Focus on Settings with Limited Resources’, with modifications made to suit local circumstances. Three surgical residents served as data collectors. All selected eligible patients were followed from hospital admission until 30 days postoperatively. Perioperative data were collected before and during surgery, using information obtained directly from patients, operation notes, anesthesia records, medical and medication charts, and direct observations. All patients were asked to provide telephone contact information (personal and/or attendant) for postdischarge follow-up calls regarding their surgical wound status. Patients/attendants received verbal and written information about signs and symptoms of SSI and wound care for the postdischarge period. Follow-up continued from the day of surgery until SSI development or the 30th postoperative day. The principal investigators provided continuous supervision of the data collection process, addressing any issues promptly.

### Study variables

**Dependent variable**: surgical site infection (SSI)

### Independent variables

**Sociodemographic factors**: Age, sex, educational status, residence area, occupational status, and marital status

**Preoperative factors**: history of previous hospitalization, preoperative hospital stay, preoperative HGB, preoperative albumin, smoking, alcohol consumption, presence of comorbidities, ASA score, and BMI

**The surgery-related factors** included previous history of surgery, type of surgery, site of operation, duration of surgery, amount of blood loss during surgery, type of anesthesia given, antibiotic prophylaxis given, timing of prophylactic antibiotics given, dose of prophylactic antibiotics, wound class, and blood transfusion given.

**Postoperative factors**: type of SSI, time at which the SSI was diagnosed, wound care given, frequency of wound care, postoperative antibiotic use, drain used, duration of drain used, and total duration of hospital stay.

### Operational definitions

**Surgical site infection (SSI)** occurs within 30 days following an operative procedure and involves skin and subcutaneous tissue, deep soft tissues of the incision (for example, facial and muscle layers) or any part of the body deeper than the facial/muscle layers that is opened or manipulated during the operative procedure AND at least one of the following (1):

A. purulent drainage from the superficial incision, from the deep incision or a drain placed into the organ/space
B. organism(s) identified from an aseptically obtained specimen from the superficial incision or subcutaneous tissue or the deep soft tissues, from fluid or tissue in the organ/space by a microbiologic testing method for clinical diagnosis or treatment
C. incision that is deliberately opened aspirated by a surgeon or physician or spontaneously dehisces and testing may or may not perform AND Patient has at least one of the following signs or symptoms: localized pain or tenderness; localized swelling; erythema; or fever (>38°C);
D. diagnosis of a surgical site infection by a surgeon or a physician
E. an abscess or other evidence of infection involving the deep incision, the organ/space detected on gross anatomical exam, histopathology exam, or imaging test

**Event:** In this research, the event of interest is the development of SSI following a particular general surgical procedure.

### Follow-up time

**Start time**: Postoperative day one (i.e., the procedure day),

**End time**: Time when SSI occurs or until postoperative day 30.

**Censored:** those patients

- Those who were referred to other hospitals during the follow-up
- Those who are nonresponsive (after four phone calls), whose phone does not work after 5 consecutive days of the trial, or who are not coming for follow-up (lost to follow-up)
- Those who did not develop SSI during the 30-day follow-up.

### Data processing and analysis

At the end of the data collection period, the collected data were exported to Stata version 14.2 (Stata Corp. LP, College Station, Texas) and then carefully cleaned, coded and analyzed.

The incidence of SSI was computed by dividing the total number of events (SSI) by the total follow-up time in person-days. Kaplan–Meier survival analysis was used to estimate the cumulative risk of developing SSI at a specific point in time and the overall median time to develop SSI. The log-rank test was used to assess the statistically significant median time difference between categories of covariates at p<0.05.

Finally, to identify independent predictors of SSI among surgical patients, a Cox proportional hazards regression model was used. First, a bivariable Cox regression model was fitted for each explanatory variable; then, those variables with a P value<0.2 in the bivariate analysis, those that were noncollinear, and those that did not violate the proportional hazard assumption were entered into the final multivariate analysis. A hazard ratio with a 95% confidence interval and p value (<0.05) was used to measure the strength of the association and to identify statistically significant predictors.

### Ethics approval and consent to participate

Ethical clearance was obtained from Wolaita Sodo University, College of Health Sciences Ethical Review Committee. An official permission letter was obtained from Wolaita Sodo University Comprehensive Specialized Hospital administration to undertake the study.

After providing written informed consent, patients who fulfilled the criteria were followed with the data collection checklist. The participants were fully informed about the details of the study and what to expect from the follow-up by the data collectors. Participation in the study was entirely voluntary, and the participants were free to ask questions and to refuse or leave at any time. It was made clear to participants that only routine clinical information would be collected and that any personal identifying information would be removed prior to extracting the data. The confidentiality of the collected information was also assured.

## RESULT

A total of 261 participants were followed and included in the final analysis. At the end of the 30^th^ follow-up period, 98 (37.5%) patients developed events of interest (SSI), and the remaining 163 (62.5%) were censored (8 patients were lost to follow-up, and 155 patients did not develop events during the 30-day follow-up period). Among those who developed the event, 70 (71.43%) were detected during the inpatient follow-up, whereas 28 (28.57%) SSIs were detected during the outpatient follow-up visit within 30 days of the follow-up period.

### Sociodemographic characteristics

The study population predominantly comprised males (67.8%) rather than females (32.2%). The participants aged over 45 years constituted the largest age group (39.8%, n=104). The majority of participants were rural residents (72.8%, n=190), with 27.2% residing in urban areas. Farmers represented the most common occupation (44.4%, n=116), followed by housewives (22.6%, n=59). Most patients were married (85.8%), and a significant proportion (42.9%, n=112) reported being unable to read and write [Table 1].

**Table 1.** Sociodemographic characteristics of patients who were admitted and operated on at the general surgery department of WSUCSH from August 1 to November 30, 2024; Wolaita Sodo Town Southern Ethiopia

| Variable | Category | Frequency (%) |
| --- | --- | --- |
| **Sex** | Male | 177(67.8) |
|  | Female | 84(32.2) |
| **Age** | 15-30 | 71(27.2) |
|  | 31-45 | 86(33.0) |
|  | >45 | 104(39.8) |
| **Residency** | Urban | 71(27.2) |
|  | Rural | 190(72.8) |
| **Occupation** | Housewife | 59(22.6) |
|  | Farmer | 116(44.4) |
|  | Government employee | 22(8.4) |
|  | Private employee | 26(10.0) |
|  | Merchant | 10(3.8) |
|  | Student | 28(10.7) |
| **Education** | College and above | 29(11.1) |
|  | Secondary school | 46(17.6) |
|  | Primary school | 67(25.7) |
|  | Able to read and write | 7(2.7) |
|  | Unable to read and write | 112(42.9) |
| **Marital** | Married | 224(85.8) |
| status | Unmarried | 37(14.2) |

### Preoperative factors and disease comorbidity

According to the present study, a minority had a history of prior hospitalization (19.9%), and a small proportion had a preoperative hospital stay exceeding 7 days (7.3%). The majority presented with preoperative hemoglobin levels >10 mg/dl (85.8%), whereas 29.5% had albumin levels ≤3.4 g/dl. Smoking (9.6%) and alcohol consumption (11.1%) were relatively infrequent. A history of comorbidity was present in 19.5% of the patients, with hypertension being the most common (70.6% of those with comorbidities). Most participants had an ASA score <3 (85.8%), and a significant proportion were underweight (BMI <18.5 kg/m², 31.0%) [Table 2].

**Table 2:** Preoperative factors and disease comorbidities among patients admitted to and operated on at the general surgery department of WSUCSH from August 1 to November 30, 2024; Wolaita Sodo Town, southern Ethiopia.

| Variable | Category | Frequency (%) |
| --- | --- | --- |
| Hx of hospitalization | Yes | 52(19.9) |
|  | No | 209(80.1) |
| Preoperative hospital stays | $\leq 7$ | 242(92.7) |
| | $>7$ | 19 (7.3) |
| Preoperative hg(mg/dl) | 7-10 | 36(14.22) |
| | $>10$ | 217(85.77) |
| Preoperative albumin | $\leq 3.4$ | 77(29.5) |
| | $>3.4$ | 184(70.5) |
| Smoking | Yes | 25(9.6) |
|  | No | 236(90.4) |
| Alcohol | Yes | 29(11.1) |
|  | No | 232(88.9) |
| Comorbidity | Yes | 51(19.5) |
|  | No | 210(80.5) |
| Types of comorbidities (n=51) | DM | 6(11.8) |
|  | Hypertension | 36(70.6) |
|  | Malignancy | 9(17.6) |
| ASA score | $<3$ | 224(85.8) |
| | $\geq 3$ | 37(14.2) |
| BMI(kg/m2) | $<18.5$ | 81(31.0) |
|  | 18.5-24.9 | 124(47.5) |
|  | 25-29.99 | 36(13.8) |
| | $>30$ | 20(7.7) |

### Surgery-related factors and medication usage patterns

Table 3 presents the surgical and medication-related characteristics of the study participants. The majority of patients underwent elective surgery (59.4%), with abdominal procedures being the most common (64.0%). Most operations lasted ≤2 hours (59.4%) with blood loss <500 ml (74.3%), and general anesthesia was commonly used (64.0%). Antimicrobial prophylaxis was administered to 85.1% of patients, with ceftriaxone being the predominant agent (72.5%) given as a single dose (83.8) within 30 minutes before incision (85.6% of those who received prophylaxis). Clean-contaminated wounds were most prevalent (49.8%). Postoperative antibiotics were prescribed to 39.8% of the patients, and drains were used in 38.7%.

**Table 3:** Surgery-related factors and medication usage patterns among patients who were admitted and operated on at the general surgery department of WSUCSH from August 1 to November 30, 2024; Wolaita Sodo Town Southern Ethiopia

| Variable | Category | Frequency (%) |
| --- | --- | --- |
| Surgery urgency | Elective | 155(59.4) |
|  | Emergency | 106(40.6) |
| Site of operation | Abdominal | 167(64.0) |
|  | Genitourinary | 51(19.5) |
|  | Thorax | 13(5.0) |
|  | Neck | 22(8.4) |
|  | Extremity | 8(3.1) |
| Duration of operation in hours | $\leq 2$ | 155(59.4) |
| | $> 2$ | 106(40.6) |
| Blood loss in ml | $< 500$ | 194(74.3) |
|  | 500-1500 | 57(21.8) |
| | $> 1500$ | 10(3.8) |
| Type of anesthesia given | General | 167(64.0) |
|  | Spinal | 94(36.0) |
| Antimicrobial prophylaxis (AMP) given? | Yes | 222(85.1) |
|  | No | 39(14.9) |
| Timing of AMP in minutes before skin incision (n=222) | $\leq 30$ | 190(85.6) |
| | $> 30$ | 32(14.4) |
| Types of AMP given (n=222) | Ceftriaxone | 161(72.5) |
|  | Ceftriaxone and metronidazole | 61(27.5) |
| Dose of AMP given (n=222) | Single dose | 186(83.8) |
|  | Multiple dose | 36(16.2) |
| Type of wound class | Clean | 92(35.2) |
|  | Clean-contaminated | 130(49.8) |
|  | Contaminated | 36(13.8) |
|  | Dirty | 3(1.1) |
| **Blood transfusion** | Yes | 44(16.9) |
|  | No | 217(83.1) |
| **History of previous surgery** | Yes | 44(16.9) |
|  | No | 217(83.1) |
| **SSI occurred?** | Yes | 98(37.5) |
|  | No | 163(62.5) |
| **Type of SSI (n=98)** | Superficial | 68(69.4) |
|  | Deep | 21(21.4) |
|  | Organ | 9(9.2) |
| **Wound care** | Yes | 104(39.8) |
|  | No | 157(60.2) |
| **Frequency of wound care given (n=104)** | Daily | 43(41.3) |
|  | Two times a day | 46(44.2) |
|  | Three times a day | 15(14.4) |
| **Drain used?** | Yes | 101(38.7) |
|  | No | 160(61.3) |
| **Duration of drain used (n=101)** | <3 days | 71(70.3) |
|  | 3-7 days | 27(26.7) |
|  | >7 days | 3(3.0) |
| **Postop antibiotic** | Yes | 104(39.8) |
|  | No | 157(60.2) |
| **Duration of hospital stay** | ≤1 week | 135(51.7) |
|  | 1-2 week | 107(41.0) |
|  | >2 week | 19(7.3) |

### Incidence of SSIs

In this study, a total of 261 surgical patients were followed for a total of 5594 person-days. During the follow-up, 98 (37.1%) of the patients developed an event (SSI), and the incidence density of SSI in the study area was 17.5 per 1000 person-days (95% CI: 14.4–21.4). Superficial SSI was the most common (69.4%), followed by deep SSI (21.4%) and organ space SSI (9.2%). Among those who developed the event, 70 (71.4%) were detected during the inpatient follow-up (before the patient was discharged from the hospital), whereas 28 (28.6%) SSIs were detected during the outpatient follow-up visit within 30 days of the follow-up period. The cumulative incidence of surgical site infection (SSI) over the 30-day postoperative period was estimated via Kaplan Meier survival analysis. The Kaplan Meier failure curve shown in Figure 1 illustrates the cumulative probability of surgical site infection (SSI) development over a 30-day postoperative period. The curve shows a rising risk of SSI, particularly within the first 10 days. By the end of the 30-day follow-up, the estimated cumulative incidence of SSI reached approximately 38%. The median time to SSI was not reached within the 30-day follow-up period, as the cumulative failure probability did not exceed 0.5 [Fig. 1].

**Fig. 1.**
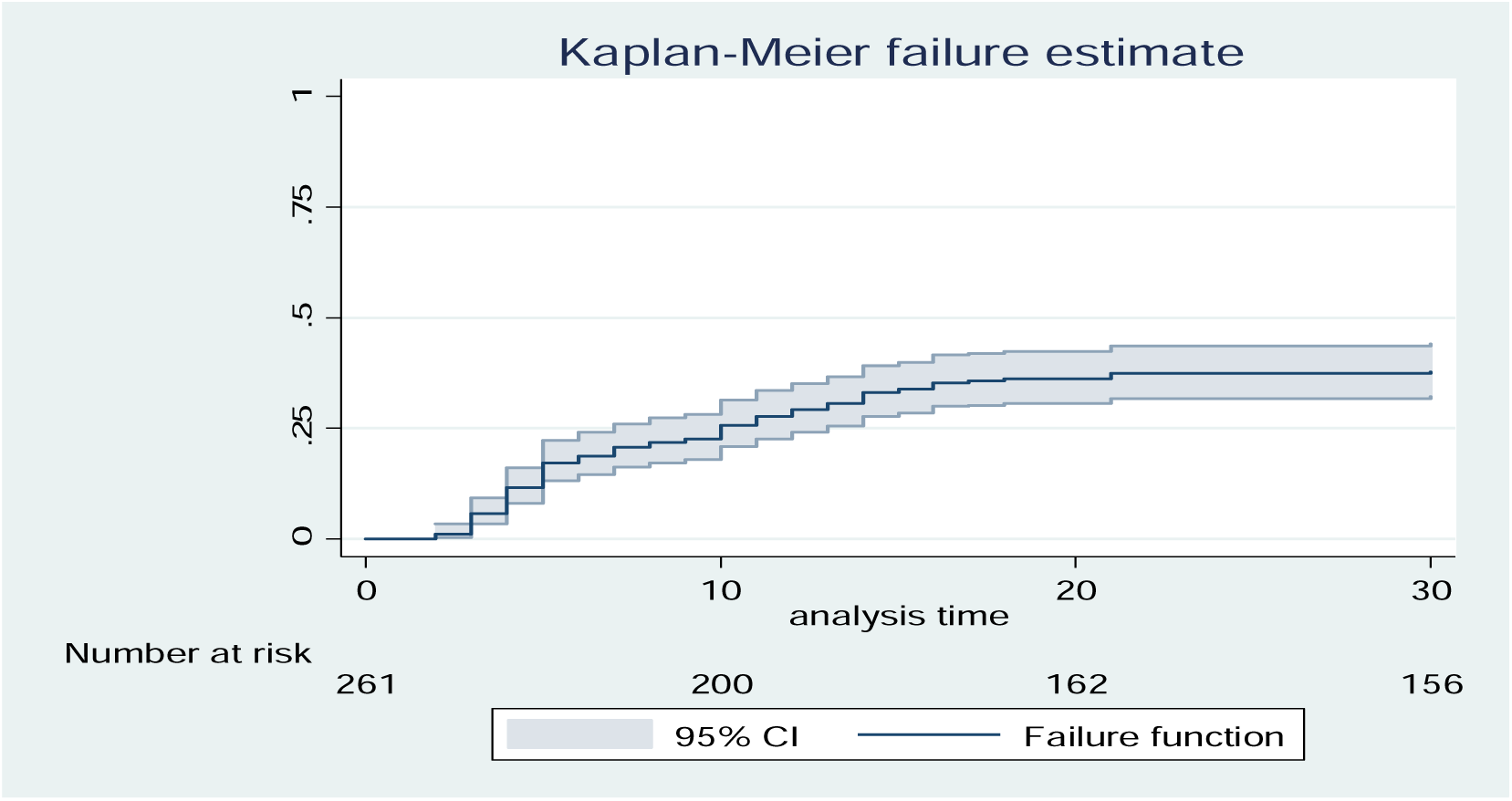
Kaplan Meier failure estimates showing the cumulative probability of surgical site infection (SSI) over 30 days after surgery among operated patients in the general surgery department WSUCSH, southern Ethiopia; 2025.

**Fig. 2.**
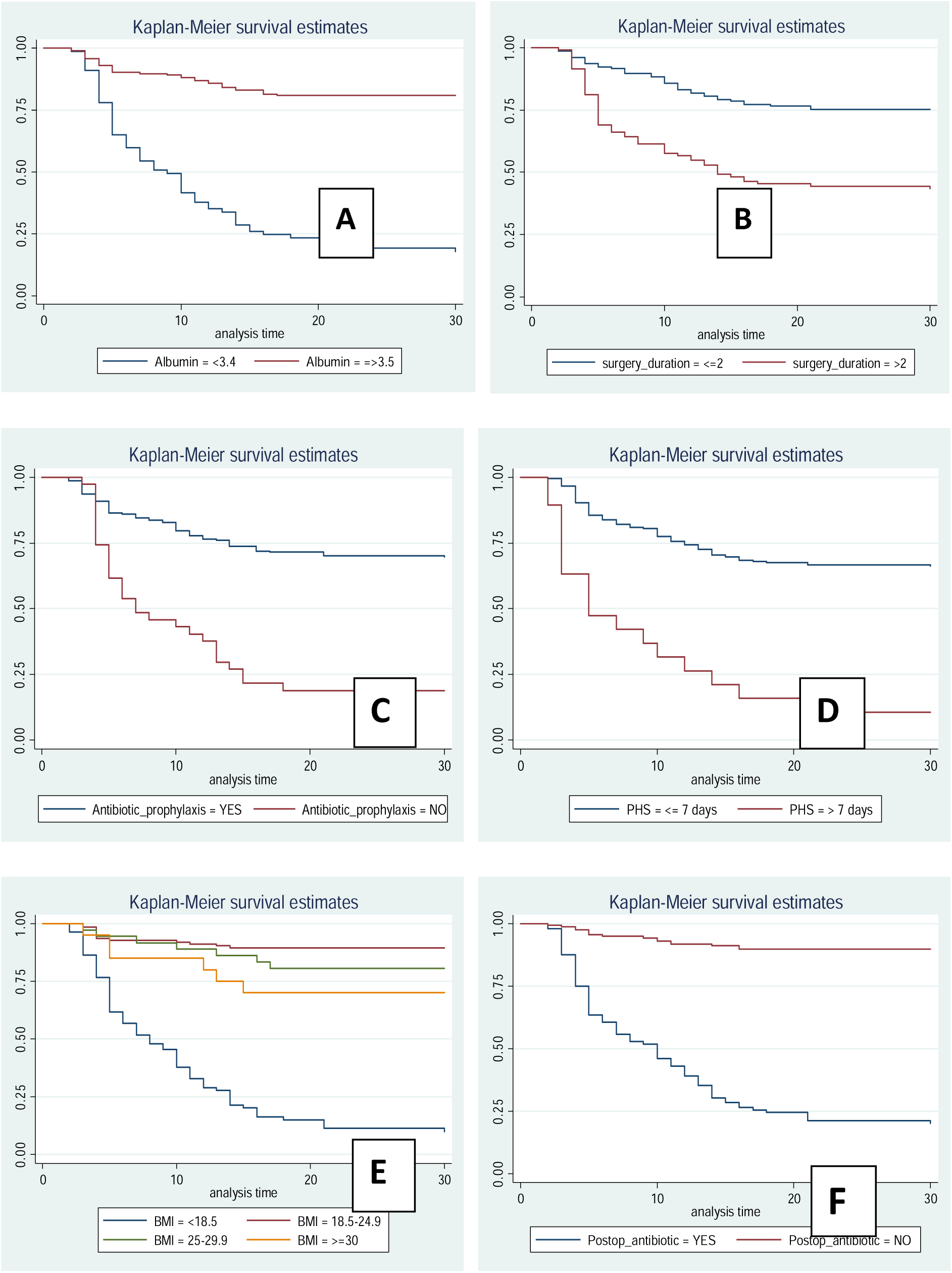
The KM survival curves that compare SSI-free survival time across different categories of (A) Pre-op albumin level, (B) duration of surgery, (C) pre-op antimicrobial prophylaxis, (D) duration of pre-op hospital stay, (E) BMI, and (F) administration of post-op antibiotics among surgically treated pateints at WSUCSH

#### Log-rank test for equality of survival functions

Log-rank test was conducted to assess the existence of any significant difference in survival time among categories of different explanatory variables. Pre-op albumin level (X2= 110.05, p=0.001), duration of surgery (X2= 30.51, p=0.001), antimicrobial prophylaxis (X2= 47.46, p=0.001), pre-op hospital stay (X2= 42.83, p=0.001), BMI (X2= 173.96, p=0.001), post-op antibiotics (X2= 147.93, p=o.001)

Kaplan-Meier survival curves were generated and a Log-rank test was conducted to evaluate significant differences in surgical site infection (SSI)-free survival time across various patient and surgical characteristics (Figure 1). The analysis revealed several factors were significantly associated with SSI-free survival. Patients with **pre-operative albumin level < 3.4 g/dL** demonstrated significantly poorer SSI-free survival compared to those with albumin ≥ 3.5 g/dL (χ2= 110.05, p=0.001). Similarly, **longer surgery duration (> 2 hours)** was associated with significantly reduced SSI-free survival compared to shorter procedures (χ2= 30.51, p=0.001). The **absence of antimicrobial prophylaxis** (χ2= 47.46, p=0.001) and a **pre-operative hospital stay exceeding 7 days** (χ2= 42.83, p=0.001) were also significantly linked to poorer SSI-free survival. Regarding **Body Mass Index (BMI)**, SSI-free survival significantly differed across categories, with underweight patients (BMI < 18.5 kg/m$^2$) demonstrating the poorest outcome (χ2= 173.96, p=0.001). Additionally, patients who **received post-operative antibiotics** exhibited significantly lower SSI-free survival compared to those who did not (χ2= 147.93, p=0.001).

### Factors associated with SSI occurrence

A Cox proportional hazards regression model was fitted to identify independent predictors of surgical site infection (SSI). The overall model demonstrated statistical significance (likelihood ratio χ2 = 214.61, p < 0.001). The multicollinearity among the variables included in the final model was assessed via a correlation matrix and found to be absent. The proportional hazards assumption (PHA) was evaluated via Schoenfeld’s residuals, yielding a nonsignificant global test (p = 0.5903, χ2 = 21.82), indicating no violation of the assumption.

In the multivariable Cox regression analysis, five factors emerged as independent predictors of SSI (p < 0.05). Patients classified as underweight (BMI <18.5 kg/m^2^) faced a 6.43-fold greater risk of developing SSI than did those with a normal BMI (18.5–24.9 kg/m^2^) at any point during follow-up [adjusted hazard ratio (AHR) = 6.43; 95% CI: 3.00, 13.78]. The risk of SSI progressively increased with increasing wound contamination, with clean-contaminated (AHR = 5.14; 95% CI: 1.12, 23.59), contaminated (AHR = 7.53; 95% CI: 2.11, 26.53), and dirty wounds (AHR = 15.51; 95% CI: 2.31, 105.83) showing 5.14, 7.53, and 15.51 times greater hazards, respectively, than clean wounds. Patients who did not receive preoperative antibiotic prophylaxis had a 2.72-fold greater risk of developing SSI than did those who did [AHR = 2.72; 95% CI: 1.17, 6.31]. Conversely, postoperative antibiotic prescription was associated with a 5.15-fold increased hazard of SSI [AHR = 5.15; 95% CI: 2.51, 10.55]. Finally, a total hospital stay duration greater than two weeks was associated with a 2.80-fold increased risk of SSI compared with stays of less than one week [AHR = 2.80; 95% CI: 1.10, 7.10] [Table 4].

**Table 4.**
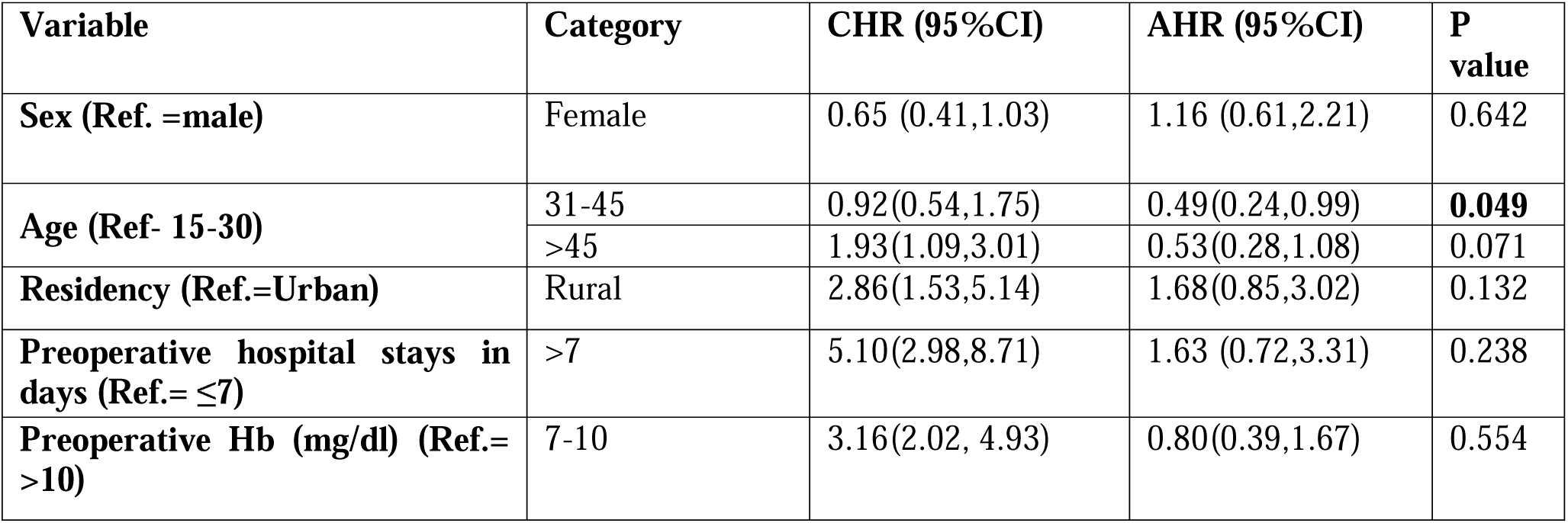
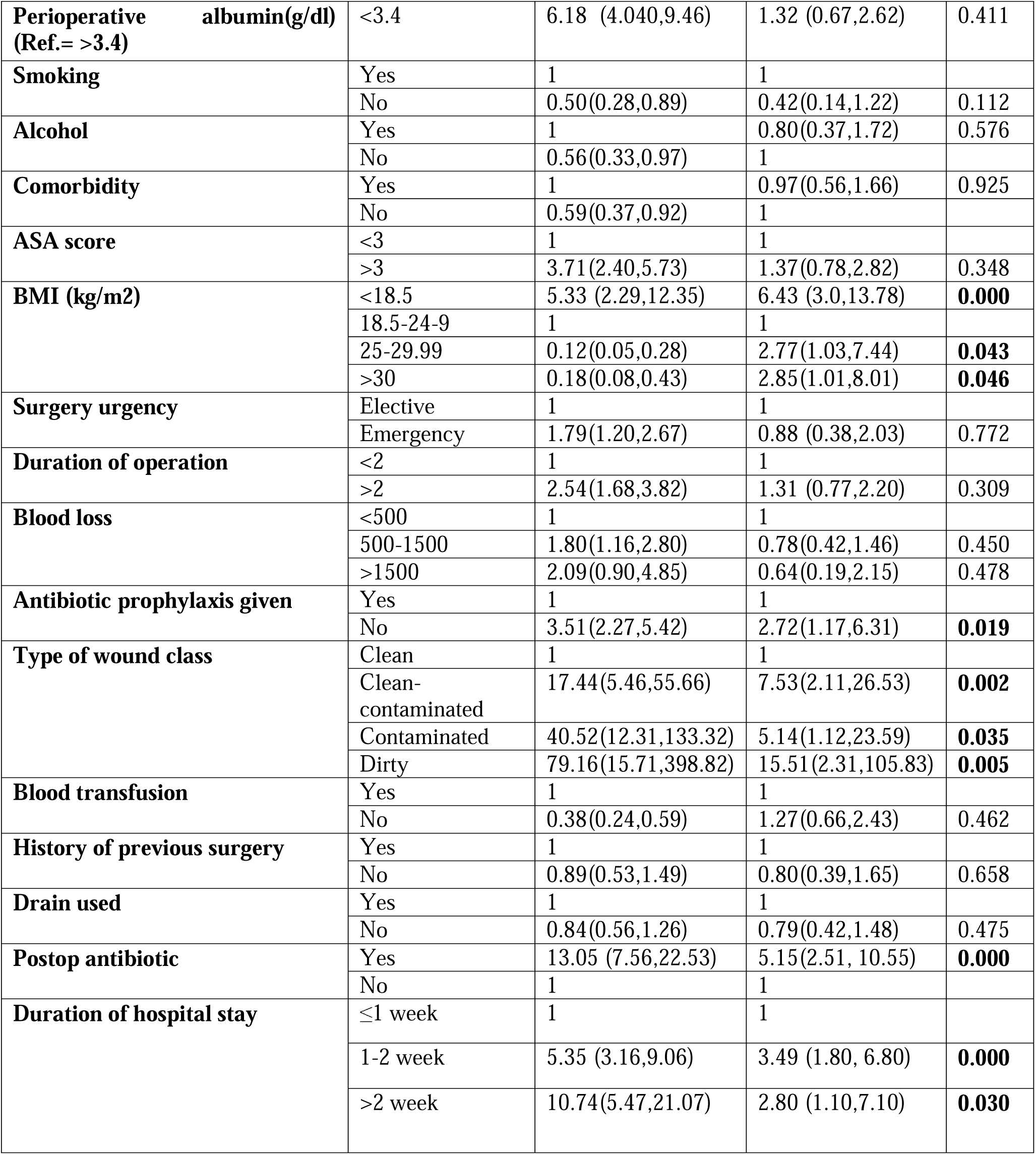
Cox regression analysis for factors associated with surgical site infections among operated patients at the general surgery department WSUCSH, southern Ethiopia; 2025.

## Discussion

This study aimed to assess the incidence and associated factors of surgical site infection (SSI) in a tertiary hospital setting in southern Ethiopia. During the 30-day follow-up period, 98 (37.5%) adult general surgical patients developed SSI, resulting in an incidence density of 17.5 per 1000 person-days. This finding is comparable to that of a prospective study conducted in the Amhara region of Ethiopia, which reported an incidence of SSI of 39.1% [15]. However, our observed incidence is notably higher than previously reported rates from various Ethiopian hospitals, including Tigray (11.5%) [13], Dessie (14.5%) [14], Jimma (21.1%) [16], Hawassa (24.6%) [17], Arba Minch (10.5%) [18], and an earlier study from Wolaita Sodo (13%) [19]. These discrepancies may be attributed to differences in infection control practices, surgical techniques, patient demographic and clinical characteristics (e.g., age, comorbidities, immunosuppression), specific study populations, and, crucially, variations in study design. Many of the earlier Ethiopian studies were retrospective or cross-sectional, relying primarily on hospital records and following patients only until discharge. This methodological approach can lead to underestimation of the true SSI burden by missing undocumented infections and those occurring postdischarge, a limitation addressed by our prospective follow-up.

Comparing our findings to high- and middle-income nations, our incidence is also substantially higher than reported rates from Brazil (3.4%) [21], China (7.5%) [22], Saudi Arabia (10.6%) [23], and India (12.6%) [24]. Similar trends are observed compared to other low-income countries in Africa, including Rwanda (8.2%) [25], Tanzania (10.9%) [26], Sierra Leone (11.5%) [27], Cameroon (12.2%) [28], Nigeria (14.5%) [29], and Uganda (16.4%) [30]. These observed differences in LMICs such as Ethiopia are likely multifactorial, potentially stemming from challenges such as limited access to necessary equipment and materials for strict aseptic conditions, suboptimal patient hygiene, increased skin colonization by pathogenic flora, late patient presentation leading to contaminated wounds, and overwhelmed emergency services owing to high patient volumes.

Our findings indicate that an increased length of hospital stay is significantly associated with an increased risk of SSI. Specifically, patients with a total hospital stay of 1–2 weeks and those who stayed longer than 2 weeks demonstrated an approximately threefold greater risk of developing SSI than patients with a hospital stay of less than one week. This finding aligns with previous studies conducted both within Ethiopia [13, 19, 20] and internationally in settings such as Brazil [21] and Saudi Arabia [23]. The biological plausibility for this association lies in the increased patient exposure to nosocomial pathogens within the healthcare environment during prolonged hospitalization. This extended exposure facilitates bacterial colonization, which can subsequently contribute to SSI development through direct contact with contaminated surfaces, healthcare personnel, or ambient air. Consequently, strategies aimed at judiciously shortening the preoperative stay and facilitating early postoperative discharge, where clinically appropriate, could mitigate the risk of SSI by reducing the exposure time.

Consistent with the literature, poor nutritional status was identified as a significant predictor of SSI. Our analysis revealed that underweight patients (BMI <18.5 kg/m^2^) had an approximately sixfold greater likelihood of developing SSI than did those with a normal BMI. This finding is supported by a meta-analysis demonstrating that malnourished patients face a greater risk of SSI postsurgery [38]. Furthermore, studies in northern Tanzania and Hawassa, Ethiopia, have similarly linked low perioperative serum ALB (< 3.4 g/dl) and general malnutrition, respectively, to adverse surgical outcomes and significantly increased SSI incidence [17, 32]. A study from Nigeria also reported a positive correlation between poor nutritional status and unfavorable surgical outcomes [29]. The underlying mechanism for this association is a compromised immune system in malnourished individuals, impairing their ability to mount an effective defense against infections. However, it is important to acknowledge a contrasting finding from Rwanda, where nutritional status (encompassing both obesity and malnutrition) was not found to be associated with an increased risk of SSI [25], suggesting that context-specific factors may influence this relationship.

Our study confirmed that wound classification is a critical predictor of SSI. Patients undergoing procedures classified as clean-contaminated, contaminated, or dirty wounds presented 5.14, 7.53, and 15.51 times greater likelihoods, respectively, of developing SSI than those with clean wounds. This finding is consistent with a study from Uganda reporting higher SSI risk in dirty and contaminated wounds [30] and another from Tanzania, where contaminated wounds had a statistically higher SSI risk than clean and clean-contaminated wounds did [25]. A systematic review in Ethiopia also revealed a more than twofold greater likelihood of SSI in clean-contaminated wounds than in clean wounds [11]. The increased risk associated with contaminated wounds is attributed primarily to the introduction of pathogens into the surgical site, compromised local host defenses, and the potential for biofilm formation [33]. Conversely, a study from Saudi Arabia reported no significant association between wound class and SSI risk [34]. Such discrepancies across studies may stem from variations in the quality of healthcare, healthcare experience, general health status of populations, and socioeconomic factors between different countries and settings.

Postoperative antibiotic prescription has also emerged as an independent risk factor for SSI. Patients who received postoperative antibiotics were approximately 5 times more likely to develop SSI than those who were not prescribed antibiotics. This counterintuitive finding aligns with a previous study from Jimma, Ethiopia, where postoperative antibiotic administration was associated with a 3.21-fold greater likelihood of SSI [16]. This association is likely indicative of reverse causation: postoperative antibiotics are often prescribed in response to a perceived higher risk of infection, a difficult or complicated surgery, or early signs suggestive of an impending or already established infection. Furthermore, inappropriate use of broad-spectrum or prolonged antibiotics can disrupt the body’s normal microbial flora, creating an ecological niche for opportunistic or resistant pathogenic microbes to proliferate and potentially cause superinfections or new infections [35]. While appropriate surgical antimicrobial prophylaxis demonstrably reduces the incidence of SSI, inappropriate use escalates the risk of antimicrobial resistance, adverse drug effects, and unnecessary costs and, paradoxically, can contribute to the development of SSI [36].

### Strengths and Limitations of the Study

This study’s primary strength lies in its prospective cohort design, which enabled direct observation and longitudinal follow-up of patients up to 30 days postdischarge. This design allowed for a more accurate assessment of SSI incidence by capturing both inpatient and postdischarge cases, thereby minimizing the underestimation prevalent in retrospective studies. Furthermore, focusing on a single tertiary hospital facilitated controlled conditions for data collection related to surgical procedures and infection control practices, providing context-specific findings directly applicable to local policies at WSUCSH. However, the single-center nature of this study limits the generalizability of the results to other hospitals or regions, as patient demographics, healthcare practices, and infection control measures can vary considerably. The potential for unmeasured confounding variables (e.g., patient adherence to postoperative care, nuances in surgical techniques) also remains. Finally, challenges in maintaining complete postdischarge follow-up could have led to data loss and potential selection bias, and resource limitations may have restricted comprehensive microbiological analyses. Future multicenter studies are recommended to validate these findings across a broader population.

### Conclusion and Recommendations

This prospective cohort study revealed a high incidence of surgical site infection (SSI) (37.5%) at Wolaita Sodo University Comprehensive Specialized Hospital in Southern Ethiopia, underscoring a significant challenge to patient safety and surgical outcomes in this setting. The key independent predictors identified include underweight BMI, increased wound contamination (clean-contaminated, contaminated, and dirty wounds), lack of preoperative antibiotic prophylaxis, prolonged hospital stay, and, counterintuitively, postoperative antibiotic administration. These findings highlight critical areas for intervention, emphasizing the urgent need for comprehensive infection prevention and control strategies. Recommendations include optimizing patient nutritional status preoperatively, meticulously adhering to aseptic surgical techniques and appropriate wound classification protocols, ensuring timely and appropriate administration of prophylactic antibiotics, and judiciously minimizing preoperative and total hospital stays. The association of postoperative antibiotics with increased SSI risk warrants further investigation, potentially indicating reverse causation and highlighting the importance of appropriate antibiotic stewardship. Given the substantial burden identified, a larger, multicenter study is crucial to validate these findings across diverse settings in Ethiopia and inform the development of national, evidence-based guidelines for SSI prevention.

## Data Availability

All data produced in the present study are available upon reasonable request to the corresponding author.

